# Characteristics, temporal trends, and outcomes of intravenous thrombolysis in stroke patients aged > 80 years in China

**DOI:** 10.1101/2024.01.11.24301186

**Authors:** Chang-sheng Li, Ying-yu Jiang, Hong-Qiu Gu, Meng Wang, Zi-mo Chen, Xin Yang, Qi Zhou, Xia Meng, Chun-juan Wang, Zi-xiao Li

**Affiliations:** Department of Neurology, Beijing Tiantan Hospital, Capital Medical University, Fengtai District, Beijing 100071, China; Department of Emergency, Taihe Hospital, Hubei University of Medicine, Shiyan 442000, China; China National Clinical Research Center for Neurological Diseases, Beijing Tiantan Hospital, Capital Medical University, Beijing 100071, China; National Center for Healthcare Quality Management in Neurological Diseases, Beijing Tiantan Hospital, Capital Medical University, Beijing 100071, China

**Keywords:** ischemic stroke, alteplase, thrombolysis, elderly, 80 year

## Abstract

**Background/Purpose:** No large cohort study has examined intravenous thrombolysis (IVT) in Chinese patients aged > 80 years.We aim to evaluate temporal trends in alteplase use, clinical characteristics, and outcomes in acute ischemic stroke (AIS) patients over 80 years of age in China.

**Methods:** Data were collected from The China Stroke Center Alliance program, which is a nationwide, multicenter, prospective registry at 1751 hospitals in 31 provinces, between January 1, 2018, and December 14, 2022.The primary outcome was a modified Rankin scale score (mRS) of 0-2 at discharge, and the secondary study outcomes were an mRS score 0-1 and independent ambulation at discharge. The safety outcomes included in-hospital mortality and symptomatic intracranial hemorrhage(sICH).

**Results:** Of 212,814 patients eligible for thrombolysis, 30,902 patients were aged > 80 years; among them, 8,673 patients (median [IQR] age, 84 [82-87] years) were treated with alteplase, 52.7% of whom were female (n=4570). The usage rate of alteplase in elderly patients increased from 22.1% in 2018 to 35.7% in 2022, while the rate among younger patients increased from 30.5% in 2018 to 43.3% in 2022. Patients treated with alteplase had better short-term functional outcomes, including mRS scores 0-2 (adjusted OR [aOR] 1.13, 95% CI 1.08-1.19, p<0.001), mRS scores 0-1 (aOR 1.14, 95% CI 1.09-1.20, p<0.001), and independent ambulation at discharge (aOR 1.27, 95% CI 1.19-1.39, p<0.001). Furthermore, there was no increased risk of in-hospital mortality (aOR 0.92, 95% CI 0.79-1.08, p=0.31). However, alteplase was associated with a higher risk of sICH (aOR 2.85, 95% CI 2.48-3.27, p<0.001).

**Conclusions:** Elderly patients receiving IVT with alteplase had better short-term functional outcomes without an increased risk of in-hospital mortality; however, elderly patients are at higher risk of developing symptomatic intracranial hemorrhage.

## Introduction

Stroke is the leading cause of death in China,^1–3^ and China has the highest estimated lifetime risk of stroke among individuals aged 25 years and above.^4^ Despite the aggressive stroke prevention strategies implemented by the Chinese government, the prevalence of stroke continues to increase with the increasing population.^3,5^ Furthermore, the prevalence of stroke increases with age. In 2019, the prevalence of ischemic stroke in patients aged 65-69 was 6.58%, while the prevalence in patients aged 80 years and above was as high as 12.81%.^6^ Importantly, acute ischemic stroke in elderly patients results in a more severe neurological deficit than stroke among younger individuals, thus producing more disabling and fatal outcomes.^7^ With the deepening of China’s aging society, China’s population aged 80 years and above is growing rapidly. The population of this age group was 26 million in 2019, and it is expected to grow to 115 million by 2050.^8^ Therefore, it is necessary to determine how to reduce the disability rate of elderly stroke patients (> 80 years old).

Intravenous thrombolysis (IVT) with alteplase is an effective treatment for acute ischemic stroke by reperfusion and reduces the disability rate associated with ischemic stroke.^9,10^ However, its application has been limited based on age. In 2013 and 2014, respectively, the American Heart Association^11^ and the Chinese Academy of Neurology^12^ no longer considered age older than 80 years as a contraindication for IVT within 3 hours from symptom onset. In 2018, the IVT time window was reported to be 4.5 hours.^13,14^ As the guidelines are being revised at both domestic and international levels, many elderly patients will be eligible for benefits. As a result, the rate of intravenous thrombolysis in elderly American patients increased significantly from 2016 to 2017 compared to 2008 to 2009.^15^ To date, there has been no large cohort study of IVT in patients aged > 80 years in China.^16,17^ In addition, trends in the rate of IVT use among elderly patients remain understudied. A complete understanding of the status and trends of IVT in elderly patients in China will help managers and doctors develop more accurate treatment strategies for elderly patients and improve the rate of IVT in elderly patients.

This study aims to explore the in-hospital outcomes in elderly patients treated with IVT with or without alteplase and describe the temporal trend of the IVT rate in patients > 80 and ≤80 years old.

## Methods

The China Stroke Center Alliance (CSCA) program is a nationwide, multicenter, voluntary, prospective registry aimed at implementing a continuous quality improvement initiative for stroke. The design and data collection plan has been described in detail previously.^18^ All secondary and tertiary hospitals in China were given access to this program. This study was conducted in accordance with the principles of the Declaration of Helsinki. Ethical approval was obtained from the ethical review board of Beijing Tiantan Hospital (ethical approval number: KY2018-061-02). The design and data collection plan has been described in detail previously. Trained hospital personnel gathered data from a web-based Patient Management Tool (Medicine Innovation Research Center). The China National Clinical Research Center for Neurological Diseases served as the center for data analysis. They also consented to examine the collective deidentified data to evaluate care quality and conduct research. The following data were collected at admission: demographic characteristics (sex, age, BMI), risk factors (smoking, drinking, hypertension, diabetes, dyslipidemia, myocardial infarction, peripheral vascular disease, prior stroke/TIA), NIHSS score at admission, stroke severity, onset-to-door time, onset-to-treatment time, door-to-needle time, dose of alteplase, medical history, hospital grade (secondary and tertiary hospitals), and hospital region (east, central, and western).

### Patient Population

This study included patients with ischemic stroke from 1,751 hospitals in the CSCA from January 1, 2018, to December 14, 2022. Patients were considered to be eligible if they had symptom onset within 4.5 h and had no absolute contraindications to thrombolysis based on the guidelines.^11^ Patients were excluded if they underwent thrombectomy, received other thrombolytic agents, underwent intra-arterial thrombolysis, or had missing information, such as NIHSS score at admission, prior mRS score, unclear key time (door-to-needle time or/and onset to treatment time), dose of alteplase, and in-hospital outcomes.

### Outcome Measurement

Five in-hospital outcomes were measured. The primary outcome was a modified Rankin scale score (mRS, range from 0 [normal] to 6 [death]) of 0-2 at discharge, and the secondary outcomes were an mRS of 0-1 and independent ambulation at discharge. Safety outcomes included in-hospital mortality and symptomatic intracranial hemorrhage (sICH) per the National Institute of Neurological Disorders and Stroke (NINDS) criterion, which is defined as any cerebral parenchymal hemorrhage confirmed by computed tomography or magnetic resonance imaging (MRI) combined with any clinical deterioration within 36 hours after treatment.^9^ Predefined subgroups based on sex, stroke severity classified by NIHSS (0-4 [mild], 5-15 [moderate], 15-20 [moderate to severe], > 20 [severe]), previous stroke or TIA, diabetes, and atrial fibrillation were used to analyze the effect of treatment on the primary outcome in elderly patients with IS.

### Statistical Analysis

Continuous variables are presented as medians, while categorical variables are described as frequencies and percentages. The chi-square test was conducted to compare categorical variables between age groups at baseline, while the Kruskal‒ Wallis test was conducted to compare continuous variables.

Multivariable regression models were used to evaluate the in-hospital outcomes in elderly patients with and without alteplase. To account for differences in baseline characteristics between the two groups, analyses were performed in a propensity-weighted population using the inverse probability-weighted method to calculate the propensity score (IPTW, adjusted model 1).^19,20^ For sensitivity analysis, a logistic model without propensity weighting explaining the clustering effect was also applied and presented (adjusted model 2). The models were adjusted for covariates, including sex, age, hospital characteristics, body mass index, risk factors (previous stroke/transient ischemic attack and atrial fibrillation), and medical history (antiplatelet agents and statins).

In the subgroup analysis, we explored the interaction of the effect of the treatment regimen on the primary outcome in specific subgroups by multivariate regression models with generalized estimating equations.

Temporal trends in IVT with alteplase between specified age groups (aged ≤80 years vs. aged >80 years) from 2018 to 2022 were assessed by Cochran-Armitage trend tests.

Missing outcome variables and variables in the univariate tables were excluded. Sensitivity analysis was performed to compare the baseline differences between populations with and without missing variables.

All P values were 2-sided, with P <.05 considered significant. Due to the large sample size, small differences may have been statistically significant but not clinically significant. Consequently, we computed absolute standardized differences to compare the characteristics of elderly patients with or without alteplase, considering an absolute standardized difference exceeding 10% as significant. All statistical analyses were performed using SAS, version 9.4 (SAS Institute Inc).

## RESULTS

### Baseline characteristics

After excluding ineligible patients, 212,814 patients with IS were included across 1,751 hospitals from January 1, 2018, to 2022 December 14, 2022. Among them, 30,902 patients were aged > 80 years, 8,673 patients received IVT with alteplase, and 22,229 patients received IVT without alteplase (Figure 1, eTable 1). Among the patients aged > 80 years (Table 1), the median age of patients in the IVT with alteplase group was 84 years (IQR 82.0-87.0), with female predominance (4570, 52.7%), and the door-to-needle time was 44.2 min (median, IQR 29.0-65.0 min).

**Figure 1.**
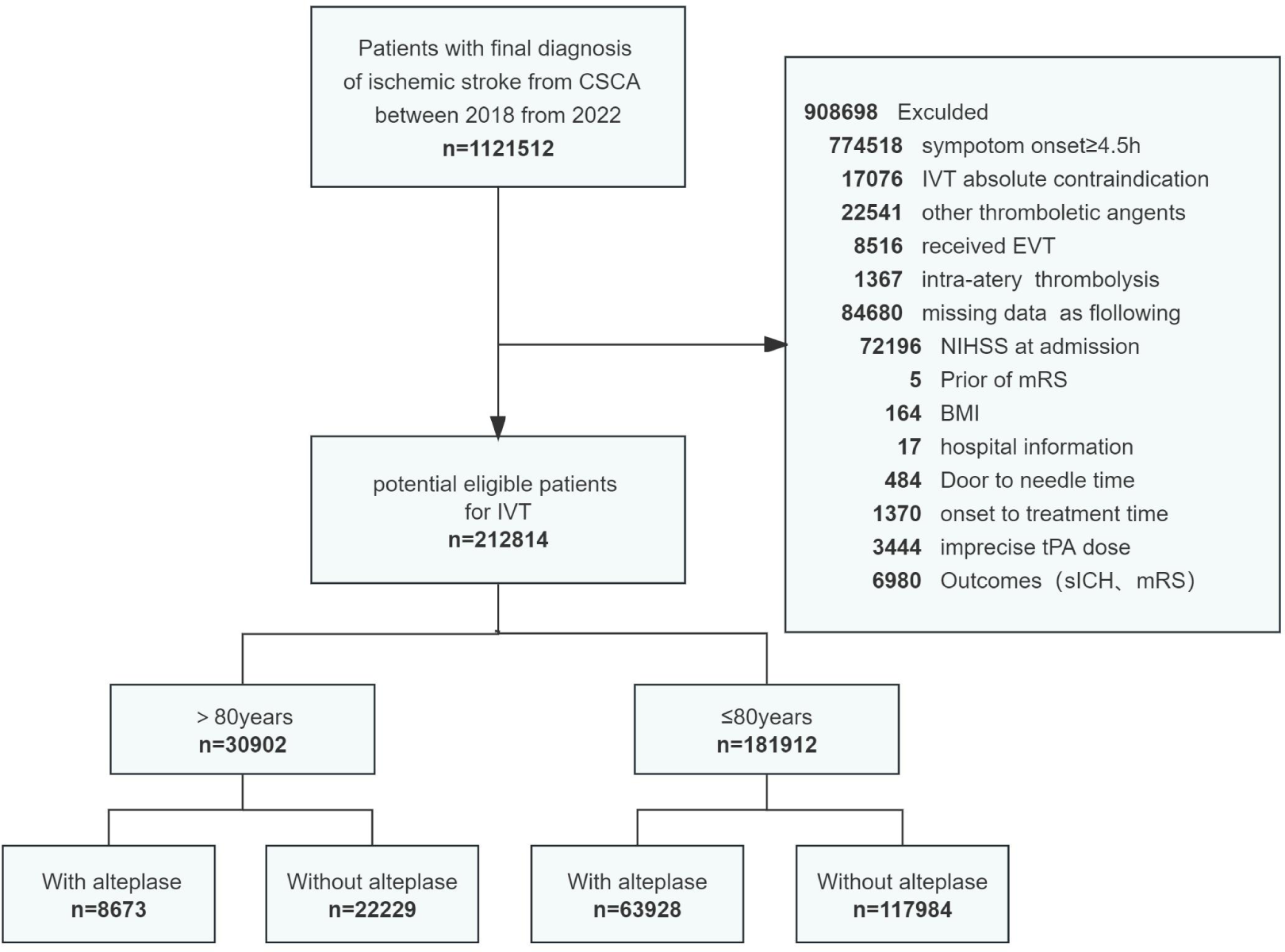
Flowchart for patient selection. BMI, body mass index; CSCA, Chinese Stroke Center Alliance; IVT, intravenous thrombolysis; NIHSS, National Institutes of Health Stroke Scale; mRS,modified Rankin Scale; tPA, tissue-type plasminogen activator

**Table 1.**
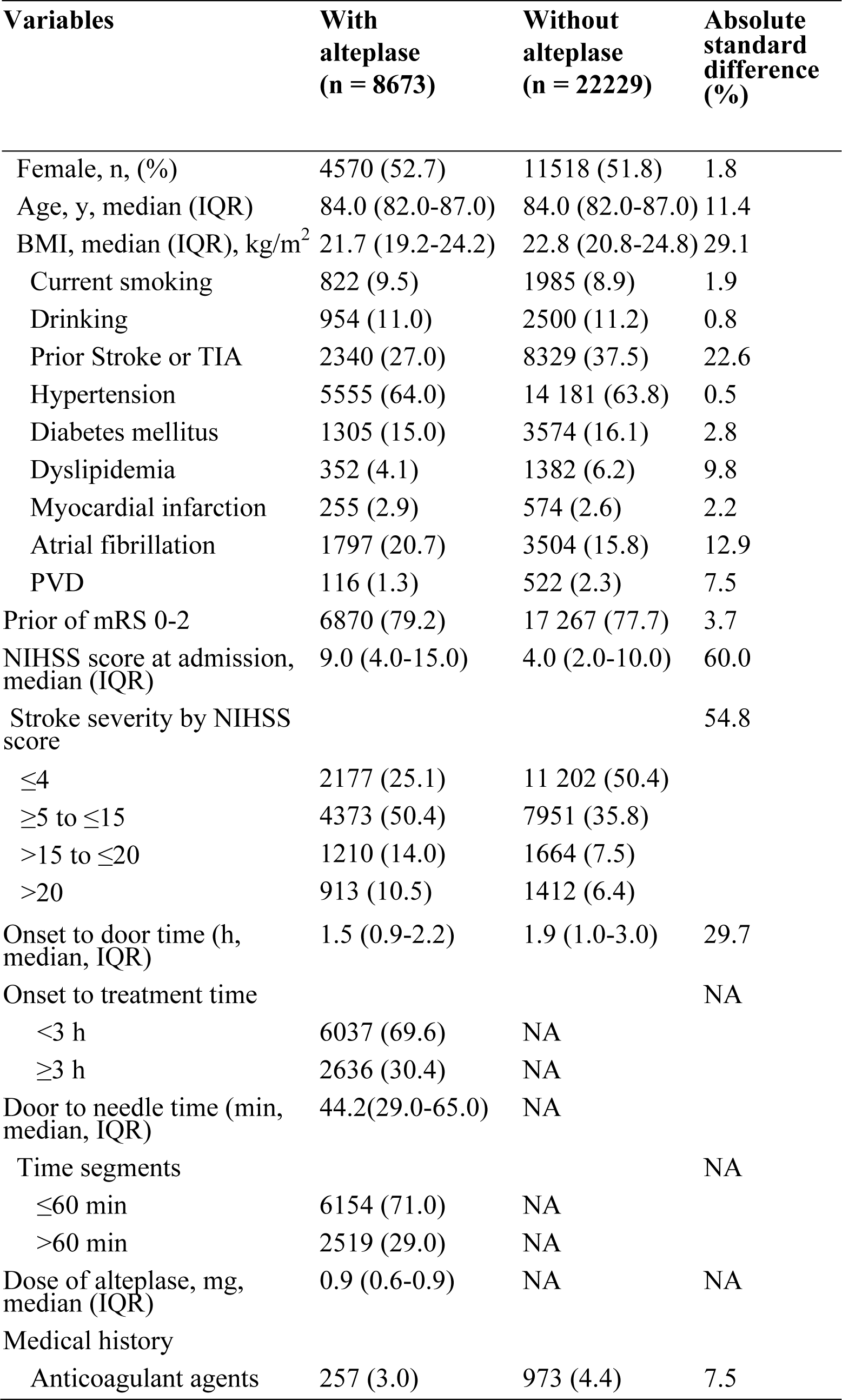
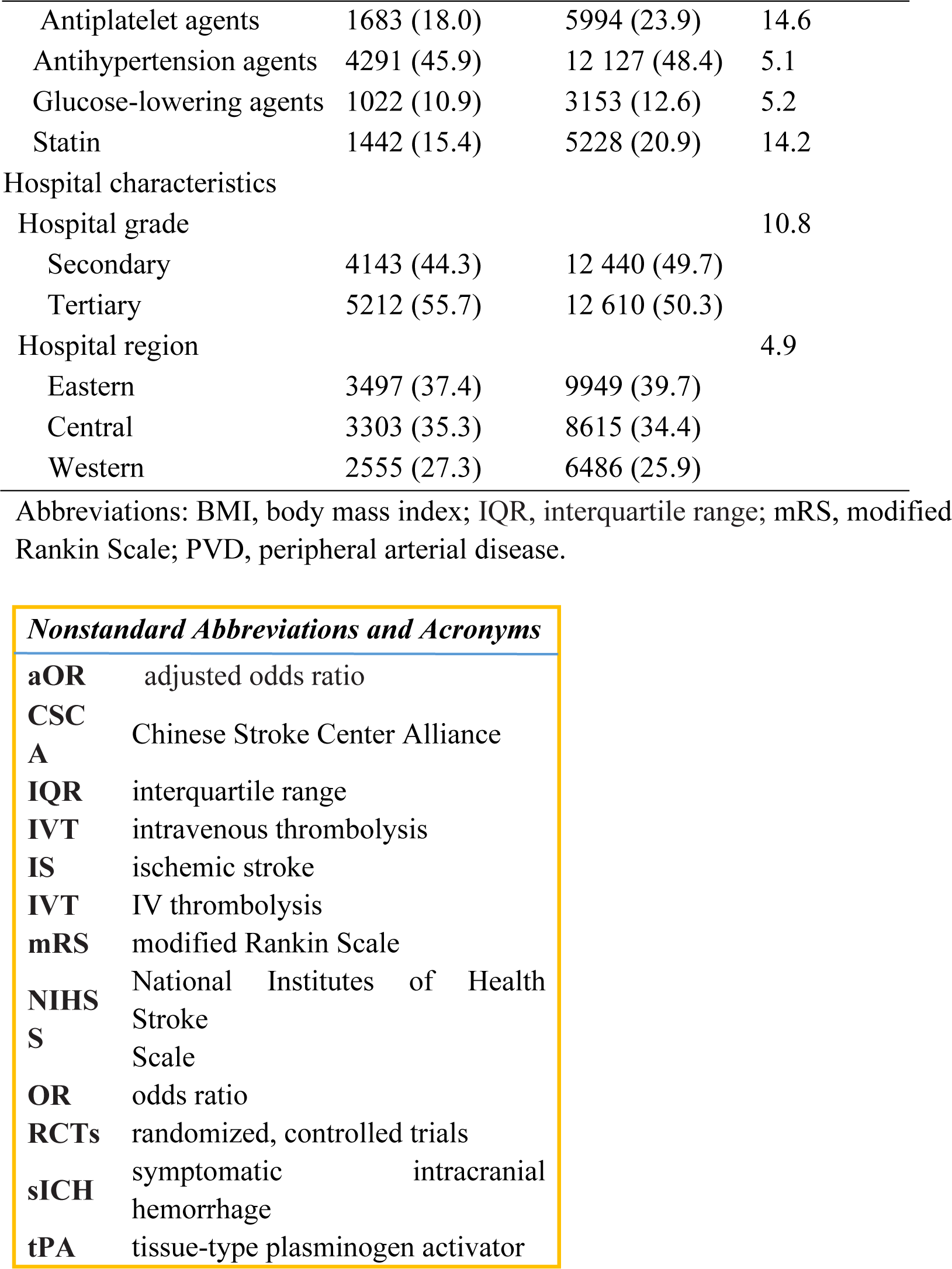
Baseline characteristics of patients aged >80 years according to assigned treatment.

| **Variables** | **With alteplase**<br>**(n = 8673)** | **Without alteplase**<br>**(n = 22229)** | **Absolute standard difference (%)** |
| --- | --- | --- | --- |
| Female, n, (%) | 4570 (52.7) | 11518 (51.8) | 1.8 |
| Age, y, median (IQR) | 84.0 (82.0-87.0) | 84.0 (82.0-87.0) | 11.4 |
| BMI, median (IQR), kg/m<sup>2</sup> | 21.7 (19.2-24.2) | 22.8 (20.8-24.8) | 29.1 |
| Current smoking | 822 (9.5) | 1985 (8.9) | 1.9 |
| Drinking | 954 (11.0) | 2500 (11.2) | 0.8 |
| Prior Stroke or TIA | 2340 (27.0) | 8329 (37.5) | 22.6 |
| Hypertension | 5555 (64.0) | 14 181 (63.8) | 0.5 |
| Diabetes mellitus | 1305 (15.0) | 3574 (16.1) | 2.8 |
| Dyslipidemia | 352 (4.1) | 1382 (6.2) | 9.8 |
| Myocardial infarction | 255 (2.9) | 574 (2.6) | 2.2 |
| Atrial fibrillation | 1797 (20.7) | 3504 (15.8) | 12.9 |
| PVD | 116 (1.3) | 522 (2.3) | 7.5 |
| Prior of mRS 0-2 | 6870 (79.2) | 17 267 (77.7) | 3.7 |
| NIHSS score at admission, median (IQR) | 9.0 (4.0-15.0) | 4.0 (2.0-10.0) | 60.0 |
| Stroke severity by NIHSS score |  |  | 54.8 |
| ≤4 | 2177 (25.1) | 11 202 (50.4) |  |
| ≥5 to ≤15 | 4373 (50.4) | 7951 (35.8) |  |
| >15 to ≤20 | 1210 (14.0) | 1664 (7.5) |  |
| >20 | 913 (10.5) | 1412 (6.4) |  |
| Onset to door time (h, median, IQR) | 1.5 (0.9-2.2) | 1.9 (1.0-3.0) | 29.7 |
| Onset to treatment time |  |  | NA |
| <3 h | 6037 (69.6) | NA |  |
| ≥3 h | 2636 (30.4) | NA |  |
| Door to needle time (min, median, IQR) | 44.2(29.0-65.0) | NA |  |
| Time segments |  |  | NA |
| ≤60 min | 6154 (71.0) | NA |  |
| >60 min | 2519 (29.0) | NA |  |
| Dose of alteplase, mg, median (IQR) | 0.9 (0.6-0.9) | NA | NA |
| Medical history |  |  |  |
| Anticoagulant agents | 257 (3.0) | 973 (4.4) | 7.5 |
| Antiplatelet agents | 1683 (18.0) | 5994 (23.9) | 14.6 |
| Antihypertension agents | 4291 (45.9) | 12 127 (48.4) | 5.1 |
| Glucose-lowering agents | 1022 (10.9) | 3153 (12.6) | 5.2 |
| Statin | 1442 (15.4) | 5228 (20.9) | 14.2 |
| Hospital characteristics |  |  |  |
| Hospital grade |  |  | 10.8 |
| Secondary | 4143 (44.3) | 12 440 (49.7) |  |
| Tertiary | 5212 (55.7) | 12 610 (50.3) |  |
| Hospital region |  |  | 4.9 |
| Eastern | 3497 (37.4) | 9949 (39.7) |  |
| Central | 3303 (35.3) | 8615 (34.4) |  |
| Western | 2555 (27.3) | 6486 (25.9) |  |

Abbreviations: BMI, body mass index; IQR, interquartile range; mRS, modified Rankin Scale; PVD, peripheral arterial disease.

***Nonstandard Abbreviations and Acronyms***
|  |  |
| --- | --- |
| **aOR** | adjusted odds ratio |
| **CSCA** | Chinese Stroke Center Alliance |
| **IQR** | interquartile range |
| **IVT** | intravenous thrombolysis |
| **IS** | ischemic stroke |
| **IVT** | IV thrombolysis |
| **mRS** | modified Rankin Scale |
| **NIHSS** | National Institutes of Health Stroke Scale |
| **OR** | odds ratio |
| **RCTs** | randomized, controlled trials |
| **sICH** | symptomatic intracranial hemorrhage |
| **tPA** | tissue-type plasminogen activator |

Compared with the patients without alteplase, patients with alteplase had more severe stroke (NIHSS [median, 9 vs. 5]), lower body mass index (median 21.7 [IQR, 19.2-24.2] vs. 22.8 [IQR, 20.8-24.8]), a higher proportion of previous atrial fibrillation (20.7% vs. 15.8%), a lower proportion of previous stroke or TIA (27.0% vs. 37.5%), shorter time from onset to door (1.5 h [IQR, 0.9-2.2] vs. 1.9 h [IQR, 1.0-3.0]), and lower use rates of antiplatelet drugs (17.9% vs. 24.2%) and statins (15.4% vs. 21.0%).

Patients with missing outcome data were excluded from the model for these outcomes. The sensitivity analysis showed no baseline differences between study patients with and without missing variables. (eTable 2).

### In-hospital outcomes for elderly patients

A comparison of the outcome measures is presented in Table 2. After adjusting for covariates by inverse probability weighted scores, elderly patients with alteplase had better functional outcomes at discharge, including proportion of mRS 0-2 (4815 [55.5%], aOR 1.14, 95% CI 1.09-1.20, p<0.001), mRS 0-1 (3743 [43.3%], aOR 1.13, 95% CI 1.08-1.19, p<0.001), and independent ambulation (5640 [65.0%], aOR 1.27, 95% CI 1.19-1.39, p<0.001). The elderly patients treated with alteplase had higher rates of symptomatic intracranial hemorrhage (4.9% [423/8673] vs. 1.2% [273/22229], aOR 2.85, 95% CI 2.48-3.27, p<0.001) but a comparable incidence of in-hospital mortality (2.3% [203/8673] vs. 1.3% [292/22229], aOR 0.92, 95% CI 0.79-1.08, p=0.31).

**Table 2.** Comparison of the in-hospital outcomes in patients aged >80 years for stroke IV with or without alteplase.

| Outcome variable | Outcome rates (%) |  | Odds ratio (95% CI) |  |  |
| --- | --- | --- | --- | --- | --- |
|  | With alteplase | Without alteplase | Unadjusted | Adjusted model 1<sup>a</sup> | Adjusted model 2<sup>b</sup> |
| **Primary outcome** |  |  |  |  |  |
| mRS 0-2 at discharge | 4815 (55.5) | 13789 (62.0) | 0.80 (0.76-0.84) | 1.14 (1.09-1.20) | 1.38 (1.30-1.47) |
| **Secondary outcomes** |  |  |  |  |  |
| mRS 0-1 at discharge | 3743 (43.3) | 10828 (48.7) | 0.76 (0.73-0.80) | 1.13 (1.08-1.19) | 1.34 (1.26-1.43) |
| Independent ambulation at discharge | 5640 (65.0) | 15935 (71.7) | 0.73(0.70-0.77) | 1.18 (1.13-1.24) | 1.27 (1.19-1.36) |
| **Safety outcomes** |  |  |  |  |  |
| sICH‡ | 423 (4.9) | 273 (1.2) | 4.12 (3.53-4.81) | 2.85 (2.48-3.27) | 3.35 (2.84-3.95) |
| In-hospital mortality | 203 (2.3) | 292 (1.3) | 1.80 (1.50-2.16) | 0.92 (0.79-1.08)<sup>c</sup> | 1.29 (1.06-1.56) |

CI, confidence interval.
<sup>a</sup>Model 1 adjusted for sex, age, BMI, prior stroke or TIA, atrial fiber/flutter, NIHSS score at admission, onset-to-door time, antiplatelet agents, statins, and hospital characteristics by inverse probability of treatment weighting score (IPTW).
<sup>b</sup>Model 2 adjusted by the logistic model accounting for the cluster effect without propensity weighting.
<sup>c</sup> p=0.31.
‡ According to the NINDS criterion.

### Subgroup analysis

As shown in Figure 2, we performed an exploratory subgroup analysis stratified by sex stroke severity by NIHSS (0-4, 5-15, 15-20, > 20), previous stroke/TIA (yes vs. no), atrial fibrillation (yes vs. no) and diabetes (yes vs. no). Using multivariate regression models, similar effects of the treatment regimen on the discharge mRS score of 0-2 were seen in all subgroups except for patients combined with diabetes (p=0.59 for interaction).

**Figure 2.**
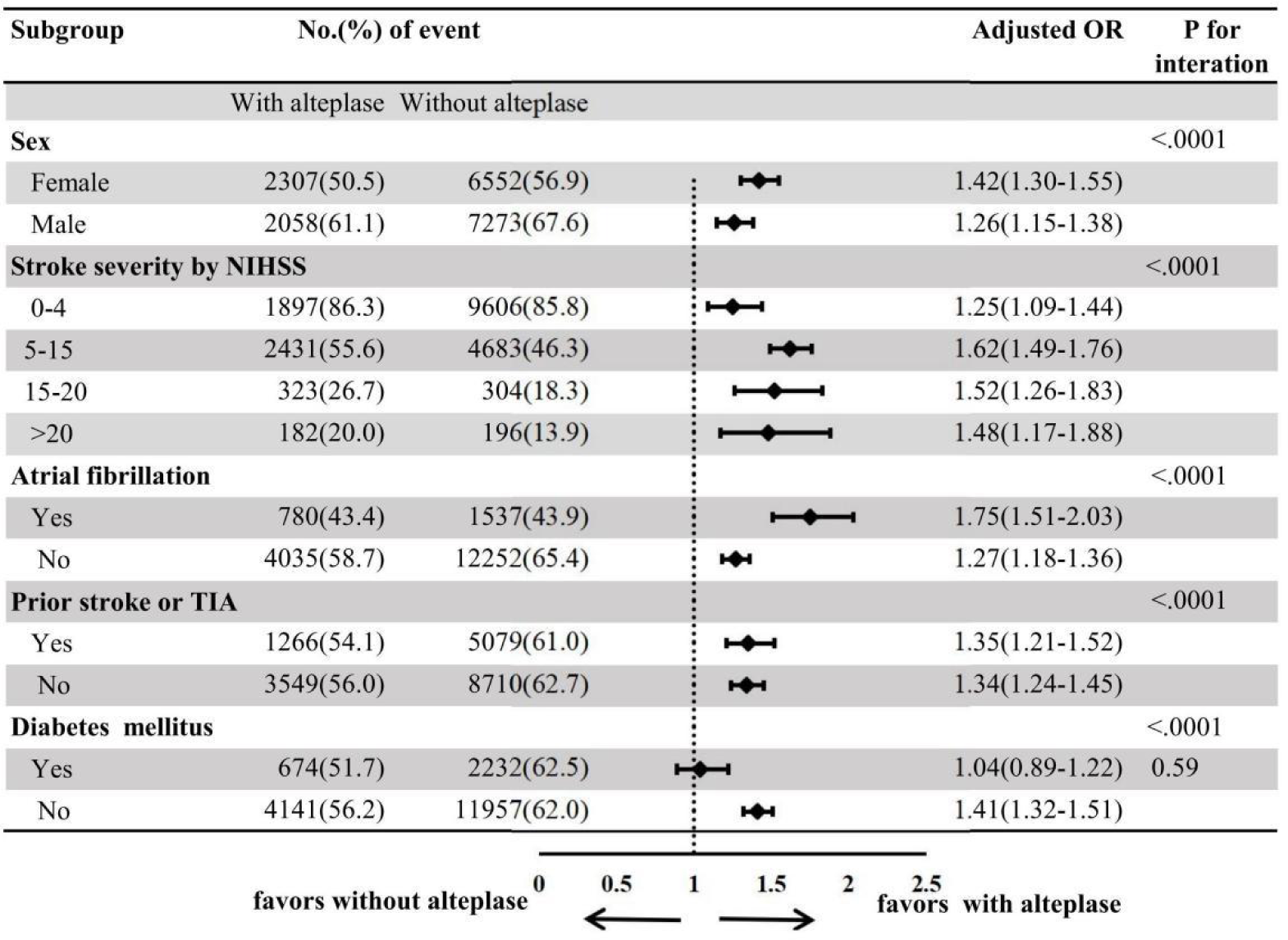
Subgroup analysis of the primary outcome in patients older than 80 years according to assigned treatment.

### Temporal trends in IVT rates among potentially eligible patients

As shown in Figure 3, the treatment rates with IVT in potentially eligible patients increased in both age groups. Throughout the study period, the use rates of patients aged > 80 years increased from 22.1% in 2018 to 35.7% in 2022, compared with younger patients from 30.5% in 2018 to 43.3% in 2022. The absolute growth rate in both age groups was similar (13.6% vs. 12.8%) throughout the study period, although patients aged older than 80 years started from a lower baseline. As a comparison, older patients show a more significant average annual growth rate of IVT compared to younger patients (10.0% vs. 7.4%).

**Figure 3.**
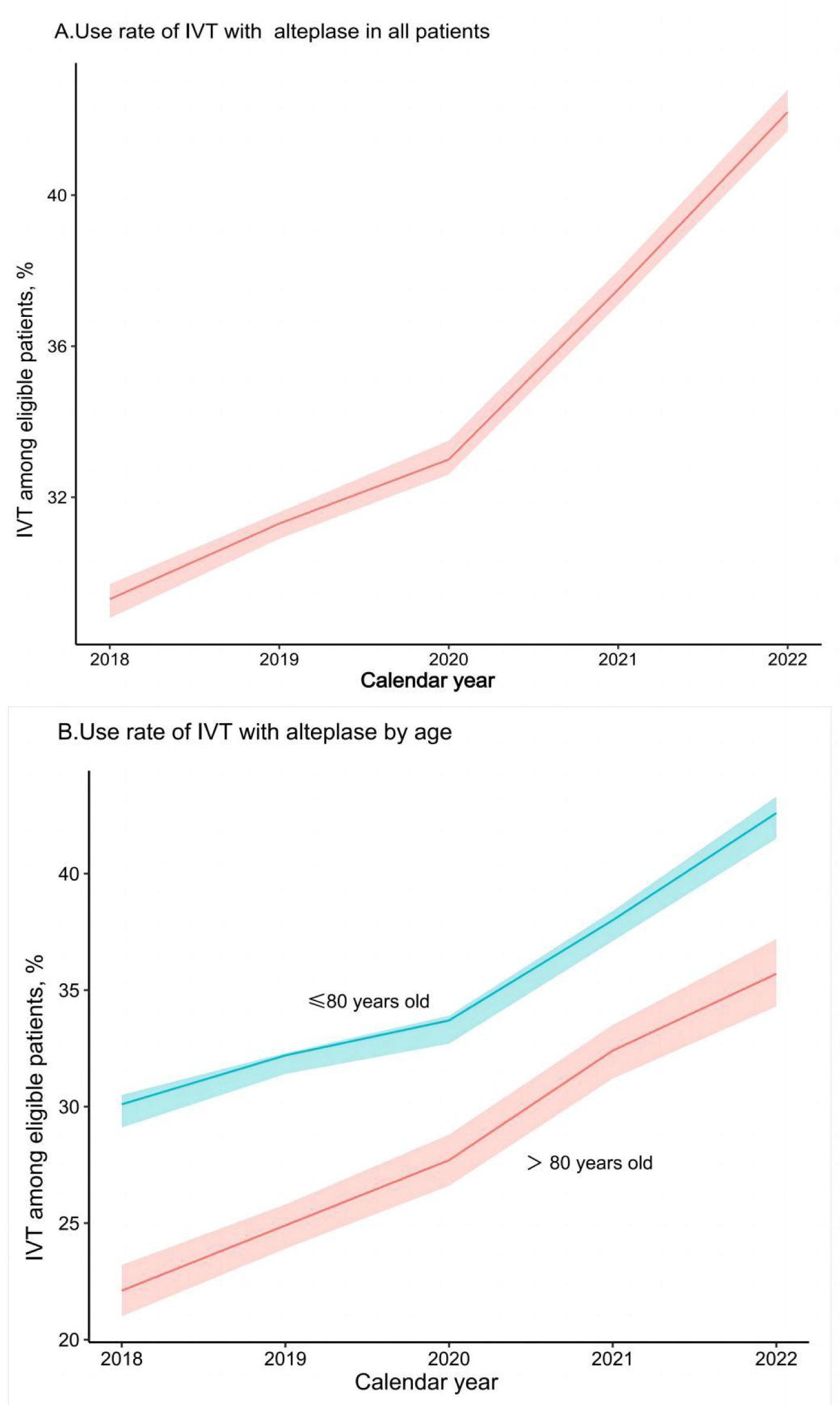
Temporal trends in rates of IV with alteplase among potentially eligible patients from 2018 to 2022. Use of IVT with in all patients (A) and in patients aged aged> 80 years and 80 years old and below(B). Shading represents 95% CIs.

## Discussion

To our knowledge, there has been no large-scale research on IVT in patients with IS aged > 80 years in China. In this large cohort study of a nationwide stroke registry, we found that treatment with IVT with alteplase in stroke patients aged >80 years was associated with better functional outcomes at discharge and did not increase the incidence of in-hospital mortality despite the increased risk of sICH compared with treatment with IVT without alteplase. Few observational studies have compared the efficacy and safety of IV thrombolysis in older patients. Our findings are consistent with the largest observational study from the Safe Implementation of Treatments in Stroke–International Stroke Thrombolysis Registry (SITS-ISTR) and the Virtual International Stroke Trials Archive (VISTA).^21^ The SITS-ISTR study consisted of 29,500 patients, with 3,472 (11.8%) of them being ≥80 years old. The results of the sensitivity analysis showed that the treatment group (IV with alteplase) was associated with higher 3-month functional outcomes than the control group (for good outcome defined as an mRS score of 0–2: OR, 2.1; 95% CI, 1.7–2.5; for excellent outcome defined as an mRS score of 0–1: OR, 1.9; 95% CI, 1.5–2.3).

Our findings revealed that the proportion of elderly patients treated with alteplase who had an mRS score of 0-2 at discharge was 55.0% (n=4815). This was higher than the proportion observed in a large study conducted in the Baden-Wuerttemberg stroke registry. In this study, the proportion of elderly patients aged 80-89 years who received IVT with mRS scores of 0-2 at discharge was 32.0% (n=2849), while for patients aged 90 years and above, it was 17.0% (n=394)^22^.

The use of thrombolysis for AIS is associated with an increased risk of sICH soon after treatment.^23,24^ Our study found that the incidence of sICH was 2.85 times higher in elderly patients who underwent IVT with alteplase than in those who did not. Nevertheless, the incidence of sICH per NINDS in elderly patients (5%) was lower than that reported in previous studies (ranging from 6.7 to 17.5%).^7,22,25,26^ This may be attributed to the lower baseline NIHSS scores in elderly patients with alteplase. The risk of symptomatic intracranial hemorrhage after thrombolysis is associated with stroke severity.^24^ In this study, the baseline NIHSS score of elderly patients with alteplase was lower than that in the RCT (mean [SD], 13.3 [6.4])^18^ and the registry study (median, 14),^7,22,27^ which could explain our low incidence of sICH. Our study found that intravenous thrombolysis did not increase short-term mortality in elderly patients with stroke. This is consistent with the results of individual analyses based on seven RCTs.^28^ This study found no significant difference in mortality within 7 days between the alteplase treatment and placebo groups (10.6% vs. 7.8%, p=0.13). Similar results were observed for death within 90 days between the two groups.

The IVT rate increased annually in older and younger patients during the study period. The average annual growth rate in older patients was higher than that in younger patients (10.0% vs. 7.4%), similar to the increasing trend in the IVT rate in developed countries. However, our research found that the IVT rate for patients within 4.5 hours of stroke symptom onset was still much lower than the 63.9% in the Get with the Guidelines-Stroke (GWTG-Stroke) in the United States.^29^ Moreover, our results showed that the rate of IVT in elderly patients was only 34.2% in 2022, which means that approximately two-thirds of eligible patients aged > 80 years did not receive IVT. Therefore, the thrombolysis rate in elderly patients requires further improvement.

Interestingly, in the subgroup analysis, we found that elderly patients with stroke and diabetes had similar short-term functional outcomes of IVT with and without alteplase. Although a history of diabetes was not an exclusion criterion for thrombolysis,^14^ it was associated with worse thrombolysis outcomes.^30,31^ Moreover, a previous study showed that patients with stroke and diabetes receiving alteplase are associated with worse in-hospital functional outcomes (mRS 0-2) than untreated patients.^32^ The benefit of IVT in patients aged > 80 years with prior stroke and concomitant diabetes mellitus is not fully established. ^14^ The recanalization rate after IVT was independently influenced by the presence of diabetes in AIS.^33^ Both chronic and acute hyperglycemia induced by diabetes may contribute to blood coagulation activation and hypofibrinolysis.^34^ Furthermore, diabetes is often associated with hyperinsulinemia, which may lead to a stronger hypercoagulability^35^ and decreased fibrinolytic activity by increased production of plasminogen activator inhibitors,^36^ and these factors reduce the vascular recanalization rate in patients with ischemic stroke. This partially explains why elderly patients in our results exhibited similar functional outcomes with thrombolysis and nonthrombolysis.

Some limitations should be mentioned before generalizing the present findings. First, our study lacked data on the lack of postdischarge 90-day outcomes. However, disability status at discharge and ambulation have been demonstrated to be closely associated with 90-day outcomes^37^; therefore, predictive factors of disability status at discharge may also predict disability changes at 90 days. Second, the design of this retrospective and nonrandomized study may lead to selection bias, and we used the inverse probability weighting method to minimize the impact of this selection bias on the results. Third, this study only included data from CSCA hospitals, not from all hospitals in China. However, the CSCA is the largest stroke registry and quality care improvement program conducted in China, with over 1 million patients with acute ischemic stroke from 1751 hospitals included in this study. A sufficiently large sample size rendered the results representative.

## CONCLUSIONS

The results of this nationwide, multicenter registry study on IVT for patients aged > 80 years with IS demonstrated that IVT with alteplase in these elderly patients is effective. Furthermore, the results revealed that higher rates of sICH do not increase in-hospital mortality. These findings add to the evidence regarding the efficacy of IVT with alteplase in elderly patients in China. The rate of IVT in elderly patients has increased annually. Nevertheless, approximately 2/3 of elderly patients do not receive thrombolysis. For elderly stroke patients with diabetes mellitus, the benefits and risks of IVT need to be carefully evaluated. Further research is necessary to determine the medium- and long-term benefits of IVT in elderly patients, especially those with diabetes.

## Data Availability

After publication of the completed study, data are available upon reasonable request, which will be considered by the corresponding authors based on the information provided by the requester regarding the study and analysis plan.

## Acknowledgments

We thank all participating hospitals, relevant clinicians, statisticians, and imaging and laboratory technicians. We would like to thank Editage (www.editage.cn) for English language editing.

